# Age acquired skewed X Chromosome Inactivation is associated with adverse health outcomes in humans

**DOI:** 10.1101/2022.04.04.22272893

**Authors:** Amy L. Roberts, Alessandro Morea, Ariella Amar, Antonino Zito, Julia S. El-Sayed Moustafa, Max Tomlinson, Ruth C. E. Bowyer, Xinyuan Zhang, Colette Christiansen, Ricardo Costeira, Claire J. Steves, Massimo Mangino, Jordana T. Bell, Chloe C.Y. Wong, Timothy J. Vyse, Kerrin S. Small

## Abstract

**Background:** Ageing is a heterogenous process characterised by cellular and molecular hallmarks, including changes to haematopoietic stem cells, and is a primary risk factor for chronic diseases. X chromosome inactivation (XCI) randomly transcriptionally silences either the maternal or paternal X in each cell of XX,46 females to balance the gene expression with XY,46 males. Age acquired XCI-skew describes the preferential inactivation of one X chromosome across a tissue, which is particularly prevalent in blood tissues of ageing females and yet its clinical consequences are unknown.

**Methods:** We assayed XCI in 1,575 females from the TwinsUK population cohort and employed prospective, cross-sectional, and intra-twin designs to characterise the relationship of XCI-skew with molecular, cellular, and organismal measures of ageing, and cardiovascular disease risk and cancer diagnosis.

**Results:** We demonstrate that XCI-skew is independent of traditional markers of biological ageing and is associated with a haematopoietic bias towards the myeloid lineage. Using an atherosclerotic cardiovascular disease risk score, which captures traditional risk factors, XCI-skew is associated with an increased cardiovascular disease risk both cross-sectionally and within XCI-skew discordant twin pairs. In a prospective 10-year follow-up study, XCI-skew is predictive of future cancer incidence.

**Conclusions:** Our study demonstrates that age acquired XCI-skew captures changes to the haematopoietic stem cell population and has clinical potential as a unique biomarker of chronic disease risk.

**Funding:** KSS acknowledges funding from the Medical Research Council (MR/M004422/1 and MR/R023131/1). JTB acknowledges funding from the ESRC (ES/N000404/1). MM acknowledges funding from the National Institute for Health Research (NIHR)-funded BioResource, Clinical Research Facility and Biomedical Research Centre based at Guy’s and St Thomas’ NHS Foundation Trust in partnership with King’s College London. TwinsUK is funded by the Wellcome Trust, Medical Research Council, European Union, Chronic Disease Research Foundation (CDRF), Zoe Global Ltd and the National Institute for Health Research (NIHR)-funded BioResource, Clinical Research Facility and Biomedical Research Centre based at Guy’s and St Thomas’ NHS Foundation Trust in partnership with King’s College London.

## Introduction

Ageing is a heterogenous process characterised by cellular and molecular hallmarks and can manifest clinically as frailty and multimorbidity(Clegg et al., 2013; López-Otín et al., 2013). Ageing is a primary risk factor for diseases such as cardiovascular disease and cancer, and a better understanding of the biomarkers of ageing promises to reduce the burden of chronic disease which significantly impacts the human healthspan(López-Otín et al., 2013).

X chromosome inactivation (XCI) evolved in placental mammals to compensate for the differences in size between the X and Y sex chromosomes. XCI transcriptionally silences either the maternal or paternal X in each cell to equalise the gene expression between XX,46 females and XY,46 males(Lyon, 1961). The selection of which X is silenced is a random process that occurs during development, with the XCI status then clonally inherited by all daughter cells. Therefore, mammalian female tissues are mosaics with respect to XCI status, with an expected ratio of 1:1. However, non-random patterns of XCI are observed, particularly in ageing individuals with up to 40% of females over 60 years old presenting preferential silencing across a tissue(Busque et al., 1996; Gale et al., 1997). This is termed age acquired XCI-skew and is highly prevalent in mitotically active blood tissue(Zito et al., 2019). XCI-skew has previously been linked to autoimmunity, which presents with a stark sex-imbalance(Chabchoub et al., 2009), as well as breast and ovarian cancers, albeit with inconsistent findings(Kristiansen et al., 2002; Lose et al., 2008; Manoukian et al., 2013; Struewing et al., 2006).

The stability of XCI-skew in blood has been demonstrated over 18-24 months and is thought to be a gradual process affecting the whole haematopoietic stem cell population rather than representing fluctuations in the active stem cell pool(Tonon et al., 1998; van Dijk et al., 2002). Therefore, though XCI-skew is a sex-specific measurable phenotype, it is a potential marker of stem cell depletion or polyclonal clonal expansion of haematopoietic stem cells, which are age-associated traits irrespective of chromosomal sex(Busque et al., 1996, 2012; Gale et al., 1997). Clonal expansion of haematopoietic stem cells is also measurable by somatic mutations shared across blood cells, indicating a common stem cell precursor(Xie et al., 2014). Clonal haematopoiesis of indeterminate potential (CHIP) is a cellular phenotype describing a pre-malignant state in which ≥4% of blood cells harbour the same somatic mutation(Jaiswal & Ebert, 2019), thus representing monoclonal expansion, and CHIP is robustly associated with all-cause mortality(Jaiswal et al., 2014), haematological cancers(Genovese et al., 2014), and cardiovascular disease(Jaiswal et al., 2017).

Given XCI-skew is potentially tagging changes to the haematopoietic stem cell pool, we hypothesised that XCI-skew may be a marker of biological ageing and a risk factor for chronic disease. We tested this hypothesis by assaying XCI-skew in 1,575 females from the TwinsUK cohort and employed prospective, cross-sectional, and intra-twin designs, to characterise the relationship of XCI with molecular, cellular, and organismal measures of ageing, and cardiovascular disease and cancer risk.

## Results

### XCI-skew is associated with chronological age and increases over the life course

We measured XCI in blood-derived DNA from 1,575 participants (median age = 61) unselected for chronic disease status from the TwinsUK population cohort(Verdi et al., 2019), which comprised of 423 MZ pairs, 257 DZ pairs, and 215 singletons. Using the normalised distribution of XCI, we derive a categorical variable in which XCI-skew equated to ≥75% XCI, and extreme XCI-skew equated to ≥91% XCI. We date-matched the XCI data with existing phenotypes from TwinsUK (e.g., blood count data, molecular markers) in the subsets of individuals on whom each phenotype was available (**Supplementary Fig. 1**).

We assessed changes in frequency of XCI-skew across increasing age and identified 12% (9 of 75) of individuals under 40 years old (yrs) displaying XCI-skew (≥75% XCI), 28% (183 of 652) of 40-59yrs; 37% (185 of 498) of 60-69yrs; and 44% (132 of 303) of those over 70yrs (**Fig. 1A**). Proportions of individuals displaying extreme XCI-skew (≥91% XCI) remains consistent at ∼3-4% below the age of 60 but increases to 7% of 60-69yrs and 9% of those over 70yrs. These results suggest a stepwise increase in prevalence of XCI-skew happening after 40 years of age, then again after 60 years of age, where we also see the first increase in prevalence of extreme XCI-skew (**Fig. 1A**). After controlling for relatedness and zygosity, we find a significant positive association between age and XCI skewing, as expected (P=2.8×10^−9^, N=1,575, **Table 1**).

**Fig. 1.**
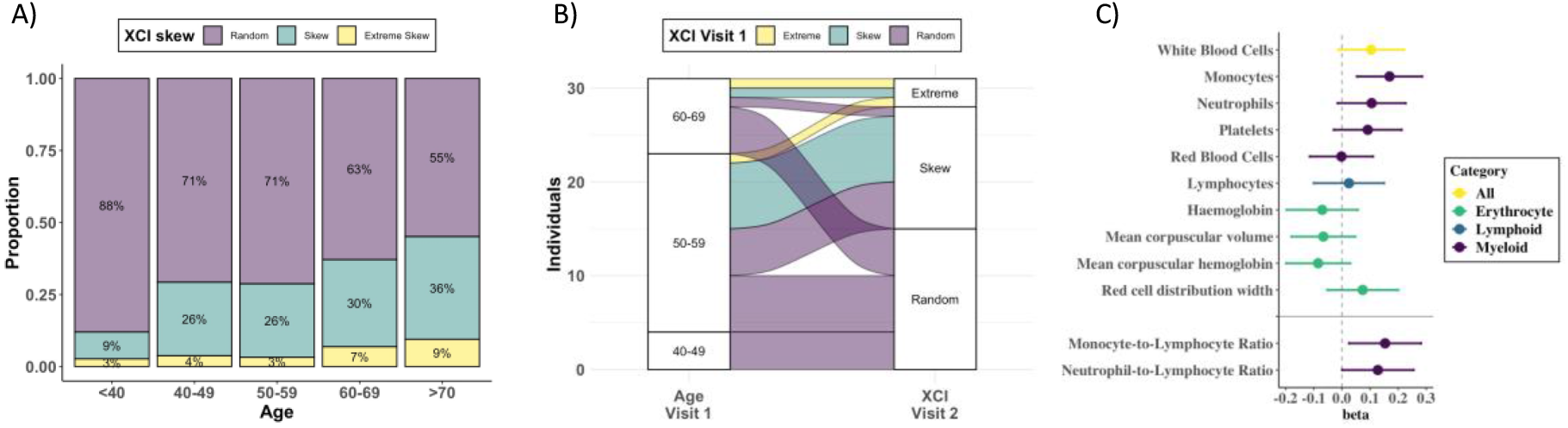
Age acquired XCI-skew across age groups and time, and associations with whole blood count data. A) Proportions of XCI-skew categories across increasing age groups. B) Sankey plot showing longitudinal changes to XCI. Colours indicate XCI at visit 1, axis 1 displays the age group of individuals at visit 1, and axis 2 displays XCI at visit 2. C) Forest plot of associations with Complete Blood Count data (top) and myeloid ratios (bottom). Measures are grouped by myeloid, lymphoid, or erythrocyte measures. Points represent the effect size of the association and lines mark the lower and upper confidence intervals.

**Table 1:** Chronological and biological age associations with XCI-skew. All data matched for year of XCI DNA sample. Bold represents significant associations after Bonferroni correction applied across 6 tests

| Phenotype | N | P-value |
| --- | --- | --- |
| Chronological Age | 1,575 | **2.8x10<sup>-9</sup>** |
| Smoking Status | 1,552 | 0.94 |
| Obesity (BMI ≥ 30) | 891 | 0.88 |
| Frailty Index | 611 | 0.59 |
| Telomere Length Shortening | 390 | 0.9 |
| GrimAge Acceleration | 137 | 0.22 |

We assessed change in XCI-skew over time using 31 individuals who had a second sample available 15-17 years prior to the main study (**Fig. 1B**. median age at visit 1 = 55.5; median age at visit 2 =72.1). The two individuals who had extreme XCI-skew at visit 1 still displayed extreme XCI-skew at visit 2. Of the eight individuals who had XCI-skew at visit 1, seven remained skewed and one progressed to extreme XCI-skew at visit 2. Of the 21 who had a random pattern of XCI at visit 1, 15 (71.4%) remained the same, and 6 (28.6%) progressed to XCI-skew at visit 2. These longitudinal data indicate that XCI-skew categorisation persists over extended periods of time and increases over the life course.

### XCI-skew is independent of known markers of biological ageing

Ageing is a heterogenous process in which an individual’s biological age can differ from their chronological age. Smoking and obesity are risk factors for accelerated ageing(Tam et al., 2020). Accelerated ageing can be estimated through measures of frailty(Clegg et al., 2013; Leng et al., 2014), and on a molecular level using measures of leukocyte telomere length (LTL) shortening(Blackburn et al., 2006) and epigenetic ageing clocks such as DNA methylation (DNAm) GrimAge(Lu et al., 2019); all these measures are associated with adverse health outcomes(Blackburn et al., 2006; Hewitt et al., 2020; Lu et al., 2019).

Given the robust association with chronological age, we sought to establish whether XCI-skew was associated with biological ageing using measures taken within one year of XCI DNA sample (**Table 1**). We observed no association with smoking status (P=0.94, N=1,552) nor obesity (P=0.88, N=891). We also found no association with LTL shortening (P=0.9, N = 390) nor DNAm GrimAge acceleration (P=0.22, N=137). Finally, we see no association with a robust frailty index (P=0.59, N=611). Together, these data, ranging from the molecular to organismal level, suggest age acquired XCI-skew is independent of many known markers of biological ageing and is potentially a unique biomarker with unexplored utility.

### XCI-skew is associated with increased monocyte abundance and decreased IL-10 levels

Changes in blood cell composition can be indicative of ill health or systemic inflammation(Kabat et al., 2017; Madjid et al., 2004; Patel et al., 2009), and a haematopoietic stem cell bias towards the myeloid lineage is observed with ageing(Pang et al., 2011). We tested for associations between XCI-skew and whole blood count data, including white cell differentials, in a subset of individuals with matched data (N=611, median age = 63). XCI-skew is association with increased monocyte abundance after multiple testing correction (P=0.0038), and we observed nominal increases in abundance across other myeloid cells (**Fig. 1B**). We next tested the hypothesis that XCI-skew was associated with a myeloid lineage bias using the Monocyte-to-Lymphocyte Ratio (MLR)(Chen et al., 2019) and Neutrophil-to-Lymphocyte Ratio (NLR)(Arbel et al., 2012), and detect an association between XCI-skew and MLR (P=0.019), and a nominal association with NLR (P=0.042) (**Fig. 1B**).

“Inflammageing” is the chronic pro-inflammatory phenotype observed in ageing and is considered an altered state of intercellular communication(López-Otín et al., 2013). Markers of inflammageing include cytokines produced by immune cells and C-reactive protein (CRP) produced by liver cells(Salminen et al., 2012). We tested for associations with serum levels of CRP and cytokines interleukin (IL)-6, IL-1B, IL-10, and TNF, in samples date-matched with the XCI DNA sample and see no association with primary markers of inflammageing (**Supplementary Table S1**). However, we observe a strong negative association with IL-10 (P=0.0008, N=27; **Supplementary Table S1**). IL-10 is a broadly expressed anti-inflammatory cytokine which can inhibit the proinflammatory responses of both innate and adaptive immune cells(Saraiva & O’Garra, 2010). Together with the blood count data which demonstrate a myeloid lineage bias, these data suggest detectable systemic changes in individuals with XCI-skew.

### Atherosclerotic cardiovascular disease risk is increased in individuals with XCI-skew

Cardiovascular disease (CVD) is the leading cause of death worldwide, and monocytes are an innate immune cell type known to be mediators in CVD disease progression and are found in atherosclerotic lesions(Libby et al., 2016). Given the association of XCI-skew with increased monocyte abundance, and the association of CHIP with CVD, we hypothesised that XCI-skew could also be associated with CVD. The atherosclerotic cardiovascular disease (ASCVD) risk score(Goff et al., 2014) (see Methods) captures traditional risk factors and gives a predicted risk of a major CVD event in the next 10 years, with an ASCVD risk score >7.5% representing intermediate risk, and an ASCVD risk score >20% representing high risk. In a cross-sectional study of 231 individuals (median age = 62) with matched health data available, XCI-skew was associated with increased ASCVD risk score after controlling for BMI and monocyte abundance (P=0.01, **Fig. 2A**). 23.5% (4 of 17) and 35.3% (6 of 17) of individuals with extreme XCI-skew have high and intermediate ASCVD risk, respectively, compared to 4.5% (7 of 155) and 21.9% (34 of 155) of individuals with random XCI.

**Fig. 2.**
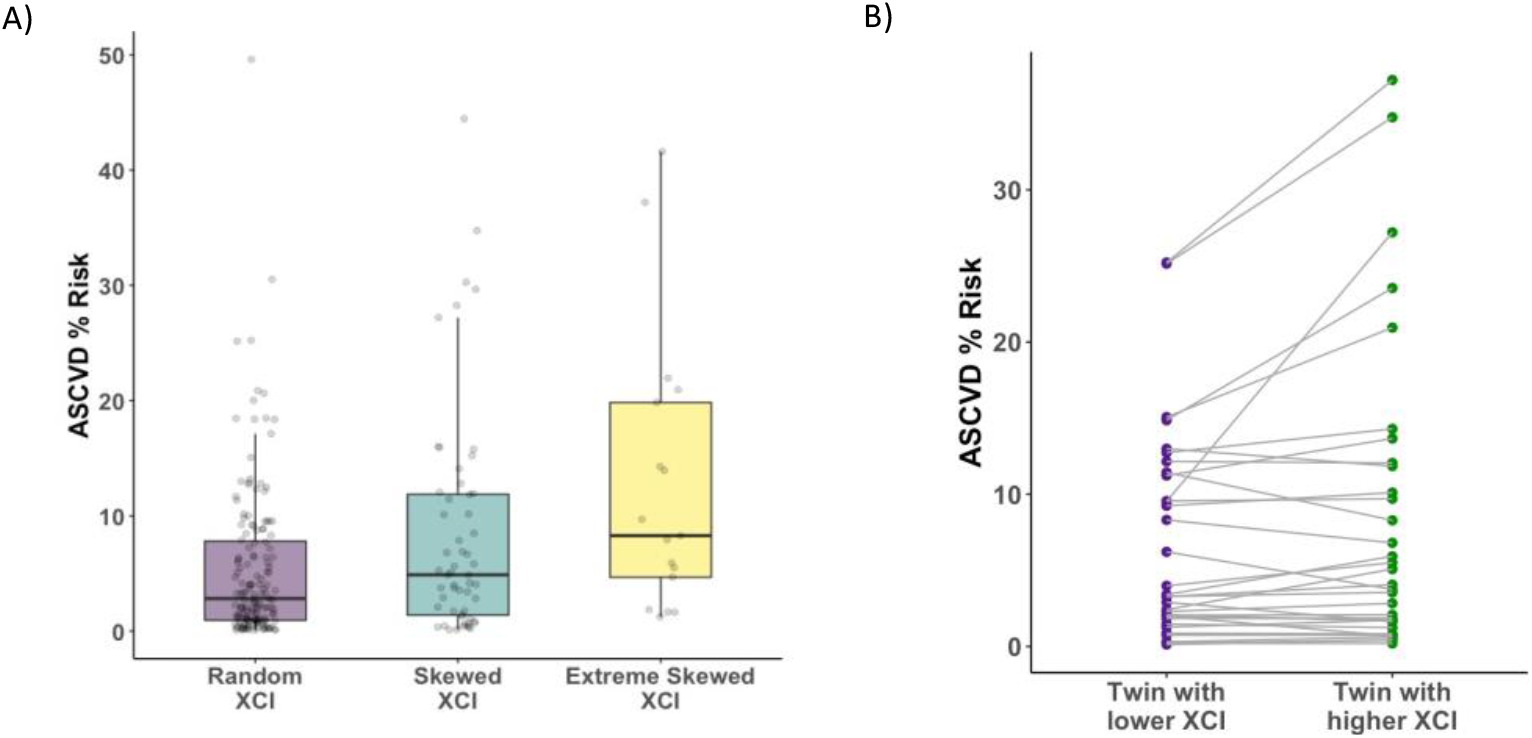
Age acquired XCI-skew and cardiovascular disease risk score. XCI-skew is associated with increased atherosclerotic cardiovascular disease risk score A) cross-sectionally after controlling for BMI and monocyte count (Linear mixed model; P=0.01), and B) within twin pairs discordant for their XCI-skew status (Wilcoxon paired t-test; P=0.025).

To ensure the observed association wasn’t spuriously driven by the age component of the ASCVD risk score (see Methods), we took age matched MZ and DZ twin pairs (n_pairs_=34) discordant for their XCI-skew status and tested intra-twin ASCVD risk scores. The intra-twin analysis validated the association between higher XCI-skew and increased ASCVD risk score (one-sided paired samples Wilcoxon test P=0.025; **Fig. 2B**), adding further support to the finding.

### XCI-skew is predictive of future cancer diagnosis in 10-year follow-up

The association of XCI-skew with cancer has largely been assessed in case-control studies focusing on cancers of female reproductive organs, with limited replicated findings(Buller et al., 1999; Kristiansen et al., 2002; Struewing et al., 2006). We conducted a prospective 10-year follow-up study (median follow-up time 5.65 years) from time of DNA sampling in 1,417 individuals (median age = 60) who were cancer-free at baseline to assess the association between XCI-skew and future cancer diagnoses (cancer events = 58; **Supplementary Table S2**). Using multivariate Cox regression analysis controlling for age, XCI-skew was associated with increased probability of cancer diagnosis (P=0.012; Hazard Ratio (HR) = 1.95 (95% Confidence Interval (CI) =1.16-3.28); **Fig. 3**).

**Fig. 3.**
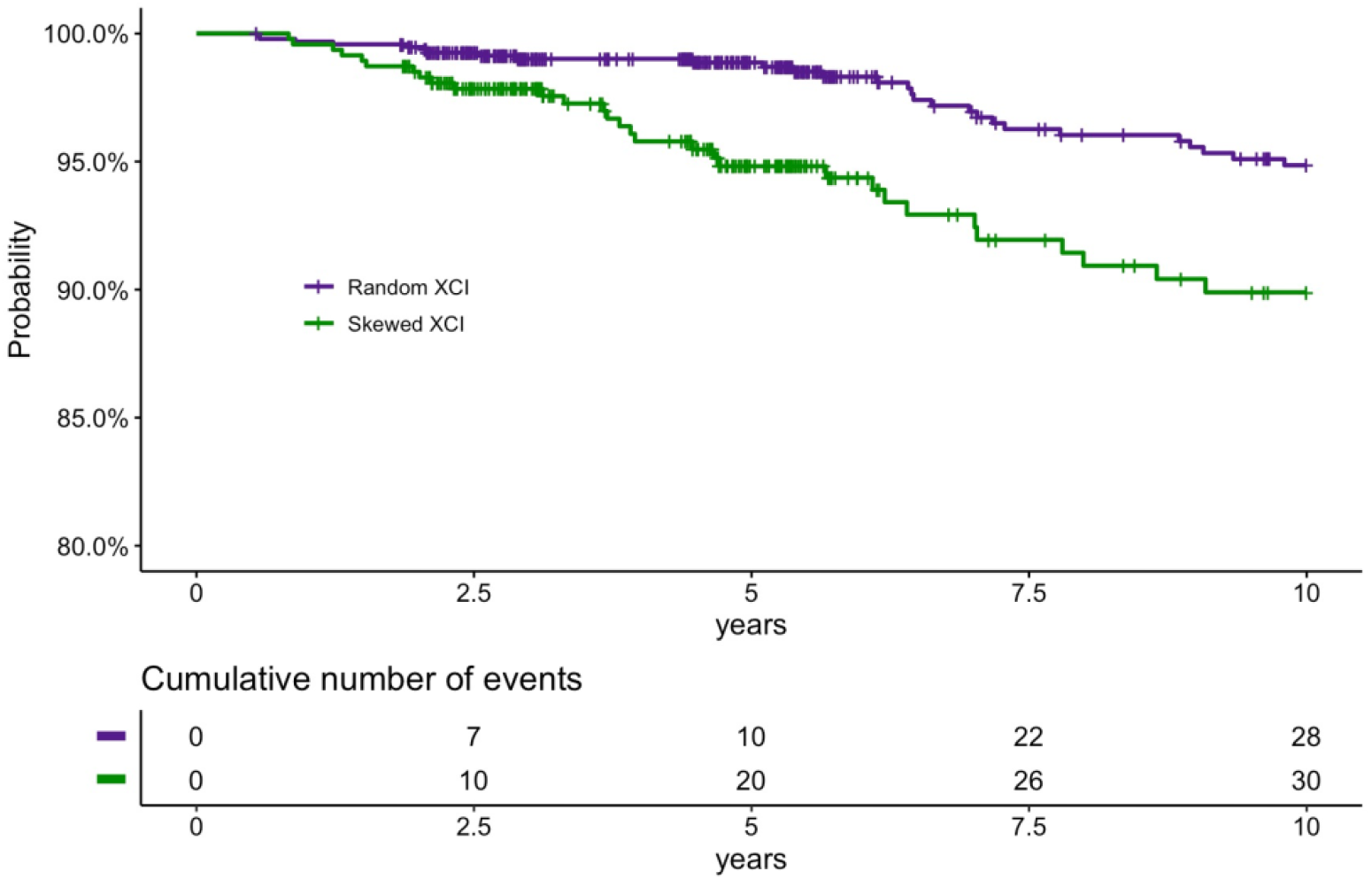
Prospective study of XCI-skew and future cancer diagnosis. Kaplan-Meier plot (top) and cumulative events (bottom) of cancer diagnosis in individuals with XCI-skew and random XCI in 10-year follow-up. XCI-skew is predictive of future cancer incidence (Cox regression, P=0.012; HR=1.95).

It has been suggested that XCI-skew could have a protective effect against all-cause mortality in cohorts selected for longevity(Mengel-From et al., 2012). However, in our 10-year follow-up study in a population cohort (median follow-up time = 5.71 years; deaths = 41), we see a non-statistically significant trend towards a positive association with all-cause mortality (P=0.29; HR = 1.39 (0.75-2.58)).

## Discussion

The age association of XCI-skew in blood tissue has long been established, with increased prevalence of females displaying XCI-skew after middle age, which is a critical time for the development of chronic disease(Busque et al., 1996; Gale et al., 1997; Zito et al., 2019). Although XCI-skew is measurable in a sex-specific manner, age acquired XCI-skew is potentially a marker of stem cell depletion or clonal expansion of haematopoietic stem cells(Busque et al., 1996, 2012; Gale et al., 1997; van Dijk et al., 2002), and could therefore have a role in age-related chronic disease, which has been robustly established for CHIP(Jaiswal & Ebert, 2019). In our cross-sectional study of 1,575 females, we identify associations of age acquired XCI-skew with an increased atherosclerotic cardiovascular disease risk score and increased probability of future cancer diagnosis.

Intriguingly, XCI-skew appears independent of other known markers of biological ageing, from molecular markers of leukocyte telomere length shortening and accelerated epigenetic ageing, to cytokine measures of inflammageing, and a robust measure of frailty, making XCI-skew a unique biomarker with clinical potential. Importantly, we also demonstrate that smoking and obesity are not risk factors for age acquired XCI-skew, and with a limited longitudinal dataset, that XCI-skew persists and increases over extended periods of time (median 16 years).

We find an association with increased probability of future cancer diagnosis in a 10-year follow-up study, with a greater risk in those 60 years or younger at study entry. Previous studies assessing the role of XCI-skew in cancer have typically used case-control studies of breast and ovarian cancers and have not been robustly replicated. The lack of replication is potentially explained by the heterogeneity in study design, including age-of-onset and therapy timing(Kristiansen et al., 2002; Struewing et al., 2006), *BRCA1* mutation stratification(Kristiansen et al., 2005; Lose et al., 2008; Manoukian et al., 2013), and threshold level for definition of XCI-skew(Buller et al., 1999). We controlled for potential confounding effects of cancer treatment on blood cells by excluding all individuals with a cancer diagnosis prior to study entry. Due to limited cancer events, our Cox regression analysis combined all individuals with XCI-skew (≥75% XCI) and extreme XCI-skew (≥91% XCI) which suggests even modest levels of skewing represents a greater risk of cancer. Follow-up studies are needed to assess the risk of cancer of specific tissues.

We also present an association with increased atherosclerotic cardiovascular disease risk score, which captures traditional risk factors and estimates the risk of developing CVD in the next 10 years(Goff et al., 2014). We were also able to utilise the powerful discordant twin design to validate this finding, thus excluding the possibility that the cross-sectional association was driven by the age component of the ASCVD risk score. Ascertaining whether XCI-skew is associated with incident CVD warrants further study.

On a cellular level, we observe that increased abundance of monocytes correlates with increased XCI**-**skew, which is of particular interest given that monocytes/macrophages are involved in the inflammatory pathophysiology of CVD(Libby et al., 2016). It is important to note however that changes in monocyte counts alone do not explain the observed levels of XCI-skew. Monocytes account for ∼10% of white blood cells whereas XCI-skew is defined here as ≥25% shift in cell mosaicism. Furthermore, age acquired XCI-skew has previously been shown across isolated neutrophils, monocytes, and T cells, with correlations between these fractions(Tonon et al., 1998; van Dijk et al., 2002). Instead, XCI-skew is likely associated with the age-related haematopoietic bias toward the myeloid lineage(Pang et al., 2011), as we see nominal associations across other myeloid cell types in addition to the monocyte- and neutrophil-to-lymphocyte ratios. Our study also demonstrates an association of reduced levels of IL-10 with increased XCI**-**skew. IL-10 is an anti-inflammatory cytokine produced by a broad range of immune cells(Saraiva & O’Garra, 2010) and subsets of monocytes differ in their capacities to secrete IL-10(Skrzeczyńska-Moncznik et al., 2008). Follow-up work on inflammatory profiles linked to XCI-skew may reveal mechanistic insights.

We derived our threshold for XCI-skew from the normalised distribution of the continuous XCI values across the cohort, and defined XCI-skew as measures 1s.d. from the mean, corresponding to XCI score ≥75%, and extreme XCI-skew as measures 2s.d. from the mean, corresponding to XCI score ≥91%. These values are very similar to thresholds used in the literature(Busque et al., 1996; Gale et al., 1997; Zito et al., 2019), but allow us to test for linear associations across the increasing thresholds of XCI-skew and demonstrates that individuals with lower levels of XCI-skew, which affects >37% of females over 60, are still at elevated risk of ASCVD and future cancer diagnosis. To avoid Type I error inflation, we did not run additional tests to compare the extreme XCI-skew group with the random XCI group.

There are some limitations to our study. Despite the high sample size of individuals with measured XCI, analyses are carried out in subsets of these individuals where date-matched phenotype data were available. In particular, the cytokine analyses have a low sample size and though we detect significant associations with IL-10, we may be underpowered to rule out the possibility of detecting a weaker association with other cytokines, and this also restricted our ability to control for cell type composition. Also, with only 41 deaths across the cohort, our all-cause mortality analysis is underpowered. A higher-powered study able to focus on specific causes of mortality is warranted. Sample sizes also limited the possibilities of running analyses on specific mortalities or cancer types, and these future studies are warranted.

In summary, we demonstrate XCI-skew is a highly prevalent cellular phenotype in females and is associated with elevated cardiovascular disease risk and predictive of future cancer incidence. Further investigations are needed to translate the biological value of XCI-skew into clinical applications for studying the association of age-related haematopoietic changes and chronic disease associations, regardless of chromosomal sex. Understanding the mechanisms underlying this phenomenon, whether XCI-skew is reflective of other ageing markers that increase disease risk, and whether it is therapeutically actionable, are areas of particular interest.

## Methods

### TwinsUK Cohort

Archival blood-derived DNA samples (collected 1997-2017) were selected from individuals of the TwinsUK population cohort(Verdi et al., 2019). Twin pairs were date matched and the final dataset of 1,575 samples comprised 423 monozygotic (MZ) twin pairs (nindividuals = 846), 257 dizygotic (DZ) twin pairs (nindividuals = 514), and 215 singletons (**Supplementary Fig. 1**). The age range of the XCI cohort is 19-99, with a median age of 61. Favourable ethical opinion was granted by the formerly known St. Thomas’ Hospital Research Ethics Committee (REC). Following restructure and merging of REC, subsequent amendments were approved by the NRES Committee London—Westminster (TwinsUK, REC ref: EC04/015, 1 November 2011).

### Human Androgen Receptor Assay (HUMARA)

XCI was measured in 2,382 samples using the HUMARA(Cutler Allen et al., 1992) method which combines methylation-sensitive restriction enzyme digest and amplification of a highly polymorphic (CAG)n repeat in the first exon of the X-linked *AR* gene, allowing for the differentiation of the active and inactive chromosomes in heterozygous individuals. 625ng of genomic DNA was divided into three aliquots and incubated for 30 minutes at 37°C with i) the methylation-sensitive enzyme *HpaII*, ii) the methylation-insensitive enzyme *MspI*, or iii) water (mock digest). The *HpaII* digest was followed by an additional 20 minutes at 80°C to avoid residual enzymatic activity. Digested DNA was amplified with FAM, VIC, NED, or VIC fluorescently labelled primers flanking the polymorphic site (Forward primer 5’-dye-GCTGTGAAGGTTGCTGTTCCTCAT-3’, Reverse primer 5’-TCCAGAATCTGTTCCAGAGCGTGC-3’). Mock and *HpaII* digested DNA were amplified in triplicate (FAM, VIC and NED), and the *MspI* digest, used as control of digestion efficiency, was amplified once (PET). To minimize technical bias and batch effects, the labelled amplified products were diluted 1:15 with nuclease-free ddH2O and pooled together with the GeneScan 500 LIZ size standard and analysed on an ABI 3730xl. Twin pairs were assayed on the same plate and plates contained a mix of both MZ and DZ pairs. Two replicates were included on each plate and a within-plate correlation of 0.99 was measured. The assay failed in 194 samples, and 601 samples were homozygous for the CAG repeat and were therefore uninformative (**Supplementary Fig. 1**).

### Calculation of XCI

Data were analysed using the Microsatellite Analysis Software available on the Thermo Fisher Cloud. The XCI status was calculated in each of the triplicates as follows:

- Allele Ratio Mock Digestion (Rm)= allele 1 peak height / allele 2 peak height
- Allele Ratio *HpaII* Digestion (Rh)= allele 1 peak height / allele 2 peak height
- Normalized Ratio (Rn)= Rh/Rm
- XCI percentage =[Rn/(Rn+1)] * 100

A coefficient of variation (CV) was calculated across the triplicates and samples with CV>0.15 were excluded from downstream analysis (n=12; **Supplementary Fig. 1**). A mean XCI percentage (0-100%) was calculated, where 50% is perfectly balanced XCI. The XCI percentage data were normalised, and absolute values of the normalised distribution were taken (XCI score) because the directionality of XCI away from 50% is uninformative (e.g., both 0% and 100% are considered equal).

### XCI-skew Categorical Variable

The categorical XCI-skew variable was created from the normalised distribution as follows: standard deviation (s.d.) <1 from the mean = random XCI (0); 1< s.d. <2 = skewed XCI (1); and s.d.>2 = extreme skew (2). As such, XCI-skew equated to ≥75% XCI, and extreme XCI-skew equated to ≥91% XCI. These thresholds are very similar to previous studies(Busque et al., 1996; Gale et al., 1997), and allowed for linear associations to be tested using the XCI-skew categorical variable.

### Statistical Analysis

In all regression models, a linear mixed effects model was used with relatedness and family structure fitted with a random intercept using the lme4 package(Bates et al., 2015). The relevant fixed effects are described in each section below and were specific to each test. All analyses were carried out using R version 4.1.1 and all plots were generated using ggplot(Wickham H, 2016).

### Chronological and biological ageing

Datasets were matched to be within 1 year of XCI DNA sample and the significance threshold after Bonferroni correction was P<0.007 to account for multiple testing across the 7 tests in this section. Chronological age was calculated at time of DNA sampling and the association was tested using XCI score as the dependent variable. Body Mass Index (BMI) measures were taken during clinical visits and obesity was defined as BMI≥30. Smoking status was classified based on longitudinal questionnaire answers(Christiansen et al., 2021). The associations were tested with XCI score as the dependent variable and obesity (obese/not obese) or smoking (ever/never smoker) as the independent variable, controlling for age as a fixed effect. A Frailty Index(Searle et al., 2008) was calculated based on longitudinal questionnaire data and used as the dependent variable, with XCI-skew as the independent variable and age and BMI as fixed effects. Leukocyte Telomere Length was measured using qPCR as previously described(Codd et al., 2010), and the normalised measures were used as the dependent variable and XCI-skew as the independent variable, with age and smoking as fixed effects. DNA methylation (DNAm) GrimAge was calculated using 450K methylation data and GrimAge epigenetic age acceleration measures were obtained from regressing epigenetic age on chronological age(Costeira et al., 2021). GrimAge Acceleration was used as the dependent variable and XCI-skew as the independent variable, with no additional covariates included in the model.

### Whole blood count data

Automated whole blood count data were date-matched to the XCI DNA sample and each of the 10 blood count variables were normalised. The significance threshold after Bonferroni correction was P<0.005 to account for multiple testing across the 10 tests. In addition, Monocyte-to-Lymphocyte Ratio (MLR) and Neutrophil-to-Lymphocyte Ratio (NLR) were calculated by dividing the total monocyte or neutrophil count, respectively, by total lymphocyte count. Associations were tested with XCI-skew as an independent variable after controlling for age, BMI, seasonality, and smoking status as fixed effects in a linear mixed effects model.

### Cytokines levels and C-reactive protein

Serum IL-1β, IL-10, IL-6, and TNF-α were measured simultaneously using the bead-based high sensitivity human cytokine kit (HSCYTO-60SK, Linco-Millipore) according to the manufacturers’ instructions. CRP concentrations from serum were measured with the Human Cardiovascular Disease Panel 2 LINCOplex Kit (HCVD2-67BK, Linco-Millipore) and with the Extracellular Protein Buffer Reagent Kit (LHB0001, Invitrogen). CRP concentrations were diluted 1:2000 prior to analysis and assayed in duplicate, as previously described(Ligthart et al., 2018). Data were normalised and associations were assessed using linear mixed models controlling for seasonality and age as fixed effects. The significance threshold after Bonferroni correction was P<0.01 to account for multiple testing across 5 tests.

### Atherosclerosis and Cardiovascular Disease (ASCVD) Risk Score

The ASCVD risk score was calculated for a subset of 231 individuals with date-matched data on age, total cholesterol, HDL cholesterol, smoking, diabetes, systolic blood pressure, and hypertension medication, as previously described(Goff et al., 2014). A linear mixed model was used to control for BMI and monocyte abundance as fixed effects(Madjid et al., 2004). Twin pairs discordant for XCI-skew but matched for date of visit and age (n=34 pairs) were used for the intra-twin study, and ASCVD risk scores were compared using a one-sided paired samples Wilcoxon test.

### Cancer and all-cause mortality

Anonymised data were obtained from the National Disease Registration Service. Study entry was the date of DNA sampling and follow-up occurred through to January 2020. For the cancer analysis, individuals who had not experienced the event were censored at 10 years, study end date, or date-of-death. All participants with a history of cancer before sampling, or within 6 months of sampling, were excluded from analyses, and reports of non-melanoma skin cancer were filtered, leaving a sample size of 1,417. For all-cause mortality, individuals who had not experienced the event were censored at 10 years or study end date. For both analyses, age and family structure and zygosity were controlled for in the Cox regression model using R package Survival(Therneau T, 2021). Proportional hazards assumptions were assessed using the cox.zph function of the Survival package. Kaplan-Meier plots were used for graphical representation of years until diagnosis. XCI-skew and extreme XCI-skew groups were combined due to the limited number of events.

## Data Availability

Data on TwinsUK participants are available to bona fide researchers under managed access due to governance and ethical constraints. Raw data should be requested via our website (https://twinsuk.ac.uk/resources-for-researchers/access-our-data/) and requests are reviewed by the TwinsUK Resource Executive Committee (TREC) regularly.

## Author Contributions

A.L.R. performed the research, analyzed and interpreted the data, performed statistical analysis, and wrote the manuscript. A.M. performed research, analyzed the data, and wrote the manuscript. A.A. performed research. A.Z. and J.S.E.M. collected and interpreted data. M.T., R.C.E.B., R.C., C.C., and X.Z. collected and analyzed data. C.J.S., M.M., and J.T.B., collected data. C.C.Y.W. and T.J.V. interpreted data. K.S.S designed the research, collected data, and wrote the manuscript.

## Acknowledgements

The authors acknowledge use of the research computing facility at King’s College London, *Rosalind* (https://rosalind.kcl.ac.uk), which is delivered in partnership with the National Institute for Health Research (NIHR) Biomedical Research Centres at South London & Maudsley and Guy’s & St. Thomas’ NHS Foundation Trusts, and part-funded by capital equipment grants from the Maudsley Charity (award 980) and Guy’s & St. Thomas’ Charity (TR130505).This work uses data that has been provided by patients and collected by the NHS as part of their care and support. The data are collated, maintained and quality assured by the National Disease Registration Service, which is part of NHS Digital.

