## Supplementary data for "Age acquired skewed X Chromosome Inactivation is associated with adverse health outcomes in humans"

Supplementary Data – *Roberts et al*

### Figures

**Figure S1**: A flowchart of sample processing and inclusion criteria for each results group

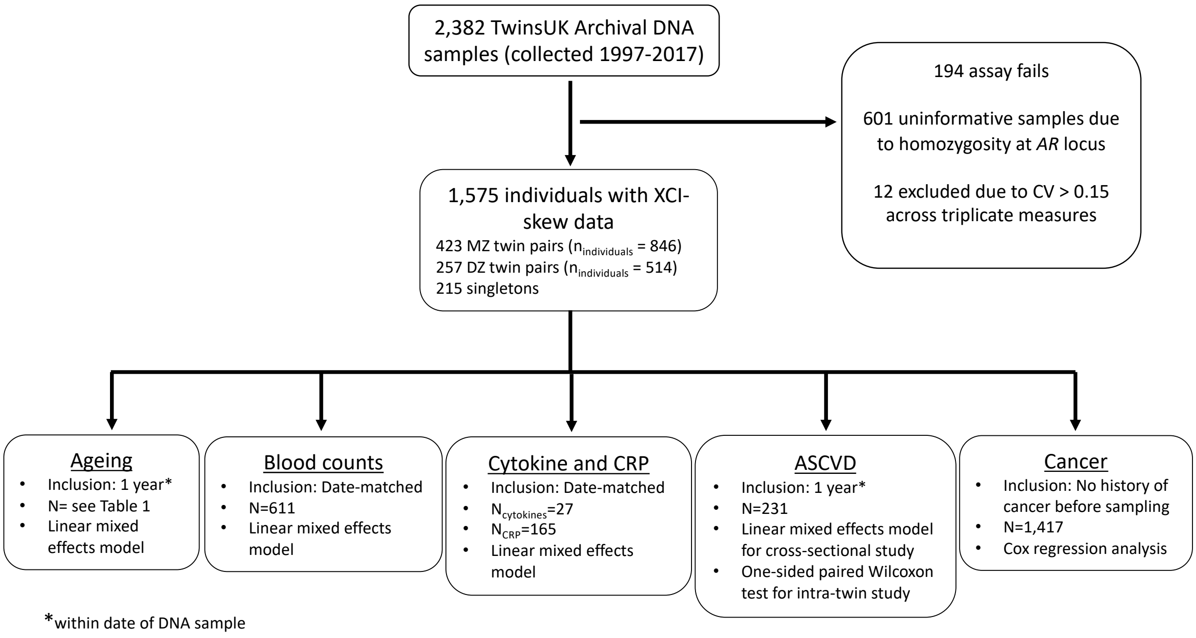


### Tables

**Table S1**: XCI-skew associations with cytokines and CRP

Age, batch effects, seasonality, and relatedness were controlled for. Bold represents significant associations after Bonferroni correction applied across 5 tests.

| Cytokine | Beta | P | N |
|---|---|---|---|
| CRP | -0.12 | 0.41 | 165 |
| IL-10 | -1.28 | **0.0008** | 27 |
| IL-1B | -1.18 | 0.02 | 27 |
| TNF | 0.28 | 0.61 | 27 |
| IL-6 | -0.35 | 0.41 | 27 |

**Table S2**: Cancer diagnoses recorded in 10-year follow-up by organ/site

| **Organ** | **Count all** | **Count under 60s** |
|---|---|---|
| Bowel | 11 | 4 |
| Breast | 26 | 10 |
| Endocrine | 1 | 0 |
| Female Reproductive Organs | 4 | 2 |
| Haematopoietic / Lymphoid Tissues | 4 | 1 |
| Oesophagus | 1 | 1 |
| Oral Cavity | 1 | 1 |
| Ovary | 4 | 1 |
| Pancreas | 1 | 0 |
| Secondary/ill-defined/Unspecified site | 1 | 0 |
| Skin melanoma | 3 | 3 |
| Urinary Tract | 1 | 0 |
| **Total** | **58** | **23** |
